# Acceptability of Overground Wearable Powered Exoskeletons for People with Spinal Cord Injury: a Multicenter Qualitative Study

**DOI:** 10.1101/2024.09.22.24313919

**Authors:** Noémie Fortin-Bédard, Alice Pellichero, Stéphanie Leplaideur, Marie-Caroline Delebecque, Caroline Charette, Willy Allègre, Alyson Champagne, Caroline Rahn, Andréanne K. Blanchette, Laurent Bouyer, Jacques Kerdraon, Marie-Eve Lamontagne, François Routhier

## Abstract

**Background:** Exoskeletons are used in rehabilitation centers for people with spinal cord injury (SCI) due to the potential benefits they offer for locomotor rehabilitation. The acceptability of exoskeletons is crucial to promote rehabilitation and to ensure a successful implementation of this technology. The objective was to explore the acceptability of overground wearable powered exoskeleton used in rehabilitation among people with SCI.

**Methods:** Fourteen individuals with SCI (9 men, mean age 47 years [14,8], majority with traumatic and thoracic lesion (T6-T12)) who had utilized an exoskeleton in Canada or in France during their rehabilitation participated in a semi-structured interview. A thematic analysis using the Theoretical Framework of Acceptability was carried out.

**Results:** Participants were motivated to use an exoskeleton during their rehabilitation. They reported several perceived benefits to its use, including better walking pattern, increased endurance and greater muscle mass. They also experienced mild pain, notable concentration demands and fatigue. Most participants reported that using exoskeletons in their rehabilitation process was appropriate and relevant to them.

**Conclusions:** Exoskeletons are generally well accepted by participants in this study. Adjustments in their use, such as conducting training sessions in obstacle-free environment and technological improvements to address the device’s restrictive characteristics, heaviness, and massiveness are however still needed.

## 1. Introduction

Over the past few years, there has been a noticeable increase in the number of people living with disabilities, especially due to the aging population [1]. Nine million people worldwide were living with a spinal cord injury (SCI) in 2019 [2]. A SCI may result in changes depending on the level and severity of the injury, such as sensory and motor impairments [3]. These impairments often cause important issues with mobility which remains a vital concern for people living with SCI and their close relatives [4,5]. In this regard, walking has been identified as one of the rehabilitation priorities for people with SCI [6,7]. Altogether, these findings highlight the need for innovative technologies supporting gait rehabilitation. As a result, lower limb powered exoskeletons were greatly developed and are now being increasingly implemented.

Exoskeletons have been recently used in facilities offering rehabilitation services for people with physical disabilities including those with SCI. Although evidence varies on the nature and extent of results, and among studies, recent studies support the safety of exoskeleton use and suggests that this technology may offer potential benefits in the rehabilitation of people with SCI [8-10]. Systematic reviews of preliminary studies found that exoskeletons allowed people with SCI to walk at modest speed [11] and engage in movements that could potentially yield health benefits [10]. More recently, a randomized trial reported an improvement in walking independence scores after participants with incomplete SCI completed 15 one-hour training sessions [9]. However, evidence of effectiveness to improve independent gait speed have not been found among SCI participants with residual walking ability [12]. In the acute phase of post-injury recovery, people with SCI reported many psychological benefits of using an exoskeleton [13].

Despite the promising effects of innovative approaches, high rates of abandonment of users in rehabilitation technology are reported notably due to failure in the implementation process [14,15]. Like other rehabilitation technologies, careful considerations should be given to the acceptability of exoskeletons by users with SCI to promote their successful implementation and sustained use in clinical settings. In that regard, Sekhon et al., 2017 have defined the acceptability of a health intervention as: “a multi-faceted construct that reflects the extent to which people delivering or receiving a healthcare intervention consider it to be appropriate, based on anticipated or experienced cognitive and emotional responses to the intervention.” [16]. Low acceptability can be a major barrier to the use of a technology and limit its widespread use [17]. Users might simply refuse or restrain activities that require the use of the technology, opting instead for more familiar activities (e.g. stretching or exercise) that align better with their usual understanding of rehabilitation.

Previous studies have investigated, sometimes as a secondary objective, the user experience of exoskeletons from the point of view of people with SCI. Various components of acceptability, such as high expectations toward this technology [18,19], considerable physical effort [13] and limited community use [20] after rehabilitation [20] have been reported. The presence or absence of these components may influence the overall acceptability of this technology. Despite its relevance for scaling up, no study has focused primarily on qualitatively exploring acceptability from the perspective of individuals with SCI [21], nor using a theoretical framework of acceptability to analyze the results. The satisfaction and their perceived acceptability of patients with SCI are critical factors in fostering their active engagement in the rehabilitation process and ensuring the successful implementation of this technology in settings that offers rehabilitation care [16,22]. This knowledge could lead to better-targeted implementation strategies. Thus, the objective of this study was to explore the acceptability of overground wearable powered exoskeleton used in rehabilitation among people with SCI.

## 2. Materials and Methods

### 2.1 Design and Participants

A qualitative study with a descriptive-interpretive approach using semi-structured interviews was conducted [23]. A qualitative design was used to provides an in-depth exploration of the acceptability of the SCI users [24]. The Consolidated criteria for Reporting Qualitative research (COREQ) checklist was used [25].

Participants were recruited from two rehabilitation facilities namely Centre intégré universitaire de santé et de services sociaux de la Capitale-Nationale (CIUSSS-CN) in Quebec, Canada and the Centre Mutualiste de Réeducation et de Réadaptation Fonctionnelles de Kerpape (CMRRFK) in Ploemeur, France. These two rehabilitation facilities offer rehabilitation care for people with SCI, which encompasses rehabilitation exoskeleton treatment among other modalities. To be eligible to participate in the interview, people with SCI had to have used an exoskeleton during their rehabilitation at the CMRRFK or the CIUSSS-CN. People who were unable to consent or complete interviews were not included. In addition, each patient was screened by a referring physician before inclusion in the study to ensures safety and relevance of the use of an exoskeleton. This study was approved by local Research Ethics Boards from the Université de Lille (2023-701-S117) and from the CIUSSS-CN (MP-13-2020-2002). Procedures were conducted in accordance with relevant laws and institutional guidelines. Participants provided their informed consent.

### 2.2. Description of Exoskeletons and Context of Use

The CIUSSS-CN uses the Indego® powered lower limb exoskeleton with functional electrical stimulation developed by Parker Hannifin Corporation® and currently supported by Ekso Bionics®. The Indego® is an exoskeleton used for people with SCI (levels C7 to L5) and for people with hemiplegia [26]. However, at the time of this study, the use of this technology was restricted to a research project at the CIUSSS-CN for future clinical implementation. Hence, participants were required to adhere to a predetermined protocol outlined within ongoing research project for the conduct of rehabilitation sessions [27].

The CMRRFK uses the hand-free and self-balancing Atalante® exoskeleton developed by Wandercraft. The use of the exoskeleton is implemented into conventional clinical therapy sessions offered to people with SCI by physiotherapists. Locomotor training with the exoskeleton is done under medical prescription.

For both center, locomotor training with the exoskeleton was done under medical prescription. The complete inclusion and exclusion criteria for the use of the Indego® and Atalante® exoskeletons are provided in Supplementary materials; Table S1 and Table S2, respectively.

### 2.2 Data collection

The recruitment of participants differed between the two rehabilitation centers, but the data collection process was similar. The recruitment began on August 20, 2022, at the CIUSSS-CN and on June 1st, 2023, at the CMRRFK. At the CIUSSS-CN, the study was introduced to individuals with SCI who had used a rehabilitation exoskeleton by a member of the research team (C.C). If they were interested in taking part in the project, a master’s student having previous experience and knowledge to conduct interviews (N.F.-B.) contacted participants by telephone or by email. The sociodemographic data of participants at the CIUSSS-CN were collected as part of the ongoing research project. At the CMRRFK, the study was presented to clinicians (N.F.-B.). People with SCI were then offered to participate in the present project by the clinicians. Sociodemographic data were collected during the interview session for participants at the CMRRFK (e.g., gender, age, exoskeleton used, time using an exoskeleton).

Semi-structured interviews were conducted in person for participants still residing in the rehabilitation center, and via telephone or videoconference using Zoom [28] for participants who had transitioned home or to in other facilities offering rehabilitation services. Interviews were conducted by two authors (N.F.-B. and A.C) who had no prior relationship with the participants. Interviews were conducted in French or in English between July 7, 2021, and July, 26, 2023 at the CIUSSS-CN and between June 5, 2023, and June 22, 2023, at the CMRRFK. All interviews were audio recorded for transcription.

An interview guide was developed by the research team members who have all together knowledge of exoskeletons and the context in which the technology is used. The interview guide (in Supplementary material) was developed with consideration to different determinants of behavioral change according to the theoretical domains framework (TDF) (e.g., knowledge, abilities, motivations and emotions toward the technology) [29] and inspired by various contextual elements that were deemed as potentially influencing the adoption and use of the exoskeleton in a larger implementation study [27]. An additional question was added to explore the satisfaction toward the exoskeleton. The interview guide included a total of 30 questions and 7 prompts.

The interview guide was not pretested considering the limited number of individuals with SCI who had used the exoskeleton, but it was validated by an expert at the CIUSSS-CN (C.R.) Additionally, the interview guide underwent validation with a physiatrist (S.L.) working with individuals with SCI at the CMRRFK, ensuring the questions were culturally relevant and appropriately adapted to the context.

### 2.3 Data analysis

Interviews were analyzed through thematic analysis [30] with a mixed approach (deductive and inductive) [31] using the conceptual Theoretical Framework of Acceptability (TFA) developed by Sekhon et al. [16] to explore the acceptability of the exoskeleton [16] (see Figure 1). An initial coding tree covering all the seven domains of the TFA (i.e., affective attitude, burden, perceived effectiveness, ethicality, intervention coherence, opportunity costs, and self-efficacy) [16] was used to analyze the interviews [16]. After becoming familiar with the interviews, two authors (N.F-B and A.P) with complementary experience (social worker and occupational therapist) independently analyzed two interviews as part of a standardization process. Then, they met to compare their interpretation of the coding tree. The first author (N.F-B) proceeded to code the remaining interviews, and meetings were scheduled to discuss the interpretation of the content. A third person, a professor-researcher (M-E.L.), was involved to provide guidance on the analysis of the interviews. The quotes included in this article have been translated from French to English using a translation software (i.e., DeepL software).

**Figure 1.**
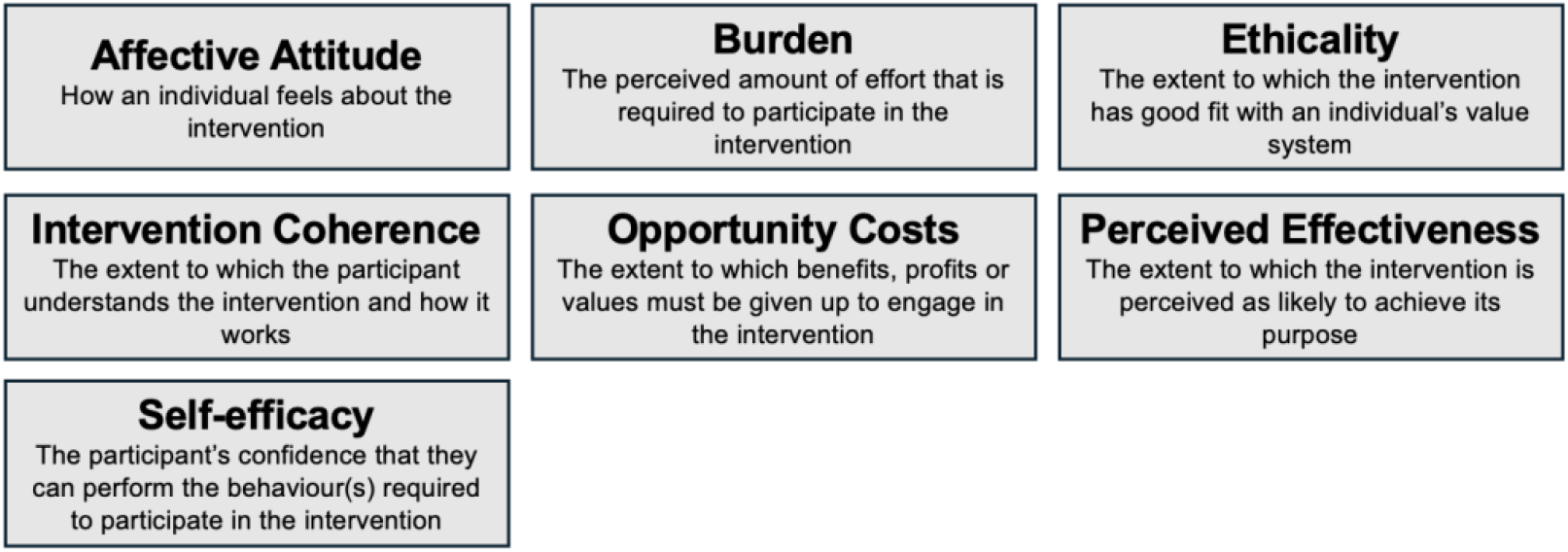
Domains of the Theoretical Framework of Acceptability (TFA) [16]

## 3. Results

Nine participants who completed the exoskeleton training program at the CIUSSS-CN, and five participants at the CMRRFK agreed to take part in the interview, for a total of 14 participants. The mean (SD) age was 46.9 (14.8) years old; most participants were men (64%) and had a traumatic SCI (79%) (Table 1). Interviews duration ranged from 19 to 63 minutes.

**Table 1.** Characteristics of the participants.

| Characteristics | n (%) |
| --- | --- |
| Age, years (mean [SD]) | 46.9 [14.8] |
| Gender |  |
| Women | 5 (36) |
| Men | 9 (64) |
| Country |  |
| France | 5 (36) |
| Canada | 9 (64) |
| Time since the injury of all participants, months (mean [SD])<sup>1</sup> | 9.9 [7.6] |
| Time since the injury of Canada's participants, months (mean [SD])<sup>2</sup> | 6.5 [3.54] |
| Time since the injury of France's participants, months (mean [SD])<sup>3</sup> | 43.8 [9.45] |
| Mechanism of injury |  |
| Non-traumatic | 3 (21) |
| Traumatic | 11 (79) |
| Self-reported level of injury |  |
| High-Cervical Nerves (C1 – C4) | 1 (7.1) |
| Low-Cervical Nerves (C5 – C8) | 2 (14.3) |
| Thoracic Nerves (T1 – T5) | 1 (7.1) |
| Thoracic Nerves (T6 – T12) | 8 (57.1) |
| Lumbar Nerves (L1 – L5) | 2 (14.3) |
| Sacral Nerves (S1 – S5) | 0 |

<sup>1</sup> Missing information for three participants.
<sup>2</sup> Missing information for two participants.
<sup>3</sup> Missing information for one participant.

Results relevant to the acceptability of exoskeletons are presented according to overall main themes including all seven domains.

### General Positive Affective Attitude

Most participants had no initial apprehension about using the exoskeleton in their rehabilitation. The exoskeleton was considered as an additional tool for achieving their rehabilitation goal. In this regard, half of the participants hoped to walk again after using the exoskeleton:

*“That’s what I was hoping for and still hope for, that it would help me walk again. [*…*] I was ready to take part in anything. That was my hope, to shock my body and then stand up most of the time. Then, the exoskeleton was part of it. We did big sessions. I wanted to take a lot of steps too, to give myself every chance*.*”* (Participant in Quebec City, Canada)

Some participants mentioned having no expectations regarding experience and outcomes to avoid disappointment. Few participants only wanted to better understand this device. Three participants reported that their expectations had been met:

*“Because it put me back on my feet, because it gave me a little hope because it gave me hope. And it [the exoskeleton] really exceeded my expectations, I was able to have fun with it*.*”* (Participant in Ploemeur, France)

Almost all participants indicated to be extremely motivated to use an exoskeleton. While walking again was identified as their main goal and motivation, others had an objective to achieve a more natural walking pattern. Their motivation to use the device stemmed from willingness to explore every available tool that could assist in their rehabilitation, as well as to contribute to the advancement of knowledge about the exoskeleton for the benefit of others. In this regard, three participants felt privileged to have had the opportunity to use an exoskeleton during their rehabilitation:

*“I was very motivated. I was told that I was lucky to be able to participate in this [to use the device], and that it wasn’t for everyone. So, well, I saw it as another chance, another tool, to help me achieve walking as normally as possible*.*”* (Participant in Quebec City, Canada)

While only one participant who considered that exoskeletons had not been utilized sufficiently for any improvement to be observed, expressed moderate satisfaction, almost all participants reported being satisfied or very satisfied with the exoskeleton, even though not all of them achieved the desired outcomes:

*“[I] am very grateful, but [I] don’t have the end result we’d like. We’d all like the miracle*.*”* (Participant in Quebec City, Canada)

A few participants reported that their relatives were excited, happy, and impressed to see the potential benefits of use on their loved ones with an SCI. However, participants had to inform their relatives about the real effects of use:

*“For them [relative], it’s magical to be able to see us again through videos of us walking vertically, especially since they mostly see us in a wheelchair. However, from their perspective, they see it more as an indication that […] we must be going to walk again. It’s always a bit more complicated to explain to them that it’s not necessarily the case, but it’s a good tool for rehabilitation*.*”* (Participant in Ploemeur, France)

### Requirements for Physical and Cognitive Engagement

Almost all participants reported using exoskeletons was physically and cognitively demanding. On the one hand, it required a high level of attention to follow instructions given by the therapist during sessions:

*“You have to be 100% focused on every step, on every breath, leaning forward, which is unnatural. You must be 100% focused. And especially at the beginning; I couldn’t talk and walk at the same time*.*”* (Participant in Quebec City, Canada)

On the other hand, the use of the exoskeleton was physically demanding, especially at the beginning of use, even if the participants were in good physical shape:

*“It’s a lot of energy, a lot of energy for a little progress*.*”* (Participant in Quebec City, Canada)

In this regard, a few individuals reported that the device was massive and heavy. A participant also expressed that the device looked like a robot. In addition, four participants reported minor soreness after or during sessions, such as pain in the back and around the lesion, tendon rubbing, spasms and small grazes. Nevertheless, participants said that the therapists had been alerted, and that the pain was minor:

*“I was more satisfied to have done it than the pain it would bring. The pain was small. [*…*] All the happiness of having done it made up for the pain it brought*.*”* (Participant in Quebec City, Canada)

Finally, one participant living outside the area where the exoskeleton sessions took place reported significant costs, particularly for travel and accommodation. Certain expenses should have been covered to enable more participants to use the exoskeleton to advance knowledge of this device:

*“Candidates from outside […] they should absolutely have an allowance for the hotel, an allowance for meals, then a travel allowance. Because if not, it [knowledge about exoskeletons] won’t advance as much*.*”* (Participant in Quebec City, Canada)

### Emerging Ethical Considerations

Few participants discussed the ethical issues regarding the use of exoskeletons. One participant reported disappointment due to the temporary effects of the use of the device: *“On the downside, there’s the disappointment, of course. When you finish, you go back into a wheelchair. That’s how it is, it’s part of the game; you must accept it. Accepting it is already a chance to be able to try it. I was aware of that*.*”* (Participant in Ploemeur, France)

Another participant expressed the possibility of false hope regarding the outcomes of the use of the exoskeleton, reminding the importance of carefully selecting potential participants:

*“These are the tests that are conducted [before use] which will determine whether you can go in or not. I think putting someone in it, especially someone on whom we are almost certain will not walk again, and giving them false hope, may not necessarily be a good idea; hence the tests conducted at the beginning*.*”* (Participant in Ploemeur, France)

Finally, a participant reported that access to the use of an exoskeleton in the rehabilitation process of people with SCI should be more inclusive:

*“Like me, I was pretty standard, no health issues, height, size, it fits, everything is fine. Granted, there are those who may not be as fortunate and might possibly need it, but in a future, maybe not too distant, we can understand that it might be enjoyable for those individuals to have access to a similar experience”* (Participant in Quebec City, Canada)

### Variable Intervention Coherence

Most participants did not have prior knowledge about exoskeletons before being approached by therapists to use the device. Even if the majority received sufficient information from their therapists, some participants reported lacking correct information on the benefits and risks of use. A participant suggested to present short videos to future participants before utilization to increase their knowledge:

*“Perhaps there could have been some small videos featuring 2-3 people without showing their faces, maybe doing the workout sequence. And, maybe showing when they put on the device. […] It would probably be interesting to be able to see a short video before signing the consent. I don’t think it scares people*.*”* (Participant in Quebec City, Canada)

One participant discovered the exoskeleton through the media

*“Like everyone else. I saw it a little in the media, but without thinking that one day I would have the opportunity to try it as a patient at the center*.*”* (Participant in Ploemeur, France)

Participants reported that individuals with an SCI having strength, endurance, and determination and who fit the inclusion criteria should have the opportunity to use an exoskeleton during rehabilitation. Some participants would have liked to use the device earlier in their rehabilitation and to continue using it for a longer period. However, one participant said that the frequency of sessions was adequate, given the effort required to participate:

*“I had two sessions a week, and then that was fine. I’m not sure I would have taken three, certainly not four, because it’s very demanding*.*”* (Participant in Quebec City, Canada)

Finally, three participants reported that they would have liked to use the exoskeleton in real-world situations, but that the usage of the exoskeleton is currently mainly in the context of rehabilitation:

*“What does it give me to take 2500 steps with two crutches if I can’t go grocery shopping? I can’t go for a walk. Because I walk on hard surfaces, so…”* (Participant in Quebec City, Canada)

### Remaining Opportunity Costs

The exoskeleton was deemed time-consuming to install, especially during the first fitting sessions. One participant reported that this initial preparation takes therapy time:

*“It’s the implementation because we have an hour of physiotherapy, and as a result, it takes three-quarters of an hour to set up. But that’s the way it is. You must*

*accept it, so you’re a guinea pig for 45 minutes for ten minutes of walking. You have to accept it*.*”* (Participant in Ploemeur, France)

A participant reported not being autonomous during the use of the exoskeleton, unlike when his wheelchair is used, resulting in a temporary loss of autonomy:

*“I could never walk as fast as the exoskeleton allows me to, but with my canes, I walk more slowly but I can walk, I can climb stairs, which I can’t do with an exoskeleton, it would limit me. I can go down a ramp, I can turn, I can*… *and then when I want to shave, and I stand up because I am able to stand, it won’t take me 15 minutes to put on the device*.*”* (Participant in Quebec City, Canada)

Finally, while most participants reported no technical problems and adequate equipment, four participants reported that the device freezes and jams, as well as problems with the waist belt, resulting in loss of time and inefficient use of therapeutic expertise.

### Positive Perception of Effectiveness

Participants’ perceptions of the effectiveness of the exoskeleton during their rehabilitation varied widely. A few participants reported benefits such as better walking pattern, increased endurance, increased muscle mass, greater leg strength, less spasticity, and improved ability to walk and walk faster:

*“I liked feeling like I did before my accident*.*”* (Participant in Quebec City, Canada)

However, one participant reported not feeling enough benefit from using this technology. In addition, few participants noted the challenge to distinguish the effects of the exoskeleton from those of other conventional rehabilitation therapies to get a clear picture of the benefits:

*“I was still willing to do anything to try to improve my condition. So, I don’t know if it improved anything, but I think it’s a combination of all the little progress I’ve had, but it’s connected to physiotherapy, the exoskeleton, and all that. That’s for sure better than lying in bed*.*”* (Participant in Quebec City, Canada)

Some potential barriers to the use of the exoskeleton were reported that can limit its use and effectiveness in rehabilitation settings, notably due to the physical environment in which the sessions took place or to get to training sessions. These included small corridors with lots of people to avoid, uneven floors and difficulty of access to sessions. However, the support received from the therapists and their knowledge, such as giving advice, being attentive to participants’ needs, and establishing a sense of trust, was reported to be an important facilitator of exoskeleton use:

*“Like being in symbiosis with the person behind you to support you. So, you need to communicate together, and it helps a lot*.*”* (Participant in Quebec City, Canada)

Overall, most participants perceived that the use of the exoskeleton during their rehabilitation was suitable and relevant for them. When questioned, almost all participants said that the use of an exoskeleton should offered to improve the rehabilitation of people with SCI:

*“I think it can be a good complement. [*…*] It’s a somewhat less medical approach to things. As I was saying, through play or through activities like this, where ultimately, one can regain a certain movement without even realizing it, to strengthen muscles, even at the level of the torso, arms, and all that*.*”* (Participant in Ploemeur, France)

### Confident Self-Efficacy

Despite the physical and cognitive demands of using the device, most participants reported no difficulties or issues in remembering the steps involved in using it. Of note, a few participants reported higher complexity in learning how to use it at first, but that the usability improved over time:

*“At first, [it’s] quite challenging. […] Because it’s not just a machine, it’s not just a robot. You must understand it. Its functioning is designed to be as close as possible to our natural movements, and you must understand that it’s a robot that’s doing it. So, you also need to*… *it accompanies you, and you must accompany it at the same time. We must become one. That’s why I talk about a second skin, because it’s a bit like that*.*”* (Participant in Ploemeur, France)

Not being too scared, having good concentration and mental focus, having realistic expectations over the outcomes of the exoskeleton, and being motivated, ambitious, determined, and persistent were qualities required to use the device according to participants:

*“You must be ready to stand up, to be supported by a machine. […] When you’ve been lying down, or at least sitting for long months and suddenly, it’s a machine that allows us to [walk] – there’s no need to be apprehensive about that, I think*.*”* (Participant in Ploemeur, France)

## 4. Discussion

The objective of this study was to explore, among individuals with SCI, the acceptability of locomotor exoskeletons as used in two rehabilitation centers located in France and Canada. Participants generally expressed positive acceptability regarding the exoskeleton used. Almost all participants reported a high level of satisfaction with the use of the exoskeleton in their rehabilitation. This satisfaction remained high despite the exoskeleton not meeting all of the participants’ expectations and a few experiencing discomforts. Most participants perceived the use of the exoskeleton as effective, experiencing little to no difficulty in its use, despite it physical and cognitive demands. While few participants reported ethical issues, some highlighted considerations regarding access to exoskeletons and possibilities of unrealistic expectations toward the use of the technology.

To support the successful implementation of exoskeletons in facilities offering rehabilitation services for people with SCI, it is crucial to have before considering others features sufficient information on the acceptability of this technology by users [32]. The results of this multi-center study, based on TFA, provide in-depth insights into the diverse experiences of individuals with SCI regarding the perceived acceptability of this technology. These results contribute to the previous studies conducted about the feasibility and effectiveness of exoskeletons used in rehabilitation. Overall, these results are consistent with a systematic literature review reporting a favorable acceptability of the use of exoskeletons among people with SCI [21]. This literature review identified the need to assess the acceptability of this technology more systematically. The use of TFA thus enables this study to respond to this knowledge limitation.

The personal characteristics of participants, including individual culture and previous knowledge, might be an important factor that influences the acceptability and use of technologies. In the present study, most participants were unfamiliar with the exoskeleton before its use, although they had been informed about the risks and benefits and their few questions have been answered. A previous study found that most people with SCI learned about the exoskeleton through traditional media channels (e.g., documentaries or news on television) [19]. The extent to which suboptimal knowledge about the effects of exoskeletons influences expectations should be examined in future studies. In this regard, clinicians involved in the study by Heinemann et al., 2018 reported that high expectations in terms of perceived benefits may be a risk for user [33]. Considering that potential benefits of exoskeleton use extend beyond physiological effects and can have a great psychological influence, these high expectations may be particularly common [34]. Consequently, future studies should focus on the development of an intervention to facilitate the adoption of realistic expectations by people using a rehabilitation exoskeleton to avoid disappointment toward technology, and, in turn, a low acceptability. This intervention could take the form of an educational activity aimed at enhancing the knowledge of both current and potential users.

Moreover, users with a SCI may feel privileged to be selected to use the exoskeleton. This feeling of privilege may bias the reported acceptability toward the exoskeleton in a favorable direction, and thus potentially influence the findings. Indeed, the limited number of exoskeletons currently available in rehabilitation centers as well as the stringent eligibility criteria (e.g., morphology, level of injury) due to the characteristics of the device are major accessibility issues [35]. This context of limited technology availability highlights the relevance of the global principles of diversity, equity, and inclusion in the use and development of innovative technologies such as exoskeletons. Consequently, as proposed in the present study, stakeholders such as developers, managers, decision makers, and others should initiate a collective reflection to ensure that exoskeletons can be used by a greater number of people with SCI or any other conditions leading to limitations in walking in the future.

The context of use of the exoskeleton [36], whether for clinical or research purpose, can also influence user acceptability. For example, in a research context, participants and clinicians must follow predetermined protocols. These protocols restrict the adaptability of exoskeletons and, to some extent, fail to consider users’ expectations, goals, and capacities. This lack of adaptability can greatly influence the acceptance of the technology, as the personalization of interventions provided is an important factor of acceptability [37]. In addition, the intrinsic features of a specific exoskeleton model that influence perceived effectiveness may also impact user acceptability [16]. For example, the Atalante exoskeleton allows users to have their arms free during its use which, in turn, allows the combination of upper-body activities with walking rehabilitation. Therefore, it is essential to analyze the goals of users when implementing a rehabilitation exoskeleton.

Limitations of our study must be considered. First, the context of use in two different rehabilitation centers with potential cultural and organizational differences, may influence the results obtained. Future studies should comprehensively assess how the context of use influences the acceptability of exoskeletons among people with SCI. Second, the time since the occurrence of the SCI was much shorter for people with SCI using an exoskeleton at the CIUSSS-CN center than at the CMRRFK center. This may influence the perceived acceptability regardless of the exoskeleton per se. In addition, the different walking prognoses among the participants could also influence the acceptability of the technology. However, we did not discern any major differences in the participants’ opinions. In addition, people with SCI at the CIUSSS-CN had to follow a research protocol for the use of the exoskeleton. The research protocol restricted the use of the exoskeleton to predefined sessions compared with more flexible sessions at the CMRRFK, where users had the possibility of performing different programs and even sporting activities. In addition, the number of usage sessions by participants could also influence their perspective. Indeed, Sekhon et al. describe well the influence of time on the acceptability of technology [16]. Third, the interview guide was based on the TDF, and consequently, users were not specifically asked about certain areas of the TFA (e.g., ethics). Finally, CIUSSS-CN participants were invited to take part in interviews only if they completed the twelve-session training program of the research protocol. Thus, the few participants who dropped out from the program were not invited to participate in the interviews. Hypothetically, these individuals could report different experiences influencing their acceptability of the technology.

## 5. Conclusions

The exoskeleton is a promising rehabilitation technology that was generally accepted by the participants in this study. Adjustments in its use, such as conducting training sessions in obstacle-free environment and technological improvements to address the device’s restrictive characteristics, heaviness, and massiveness are however still needed. In future studies, a deeper exploration of the ethical considerations will be important, along with efforts to increase the inclusivity of exoskeleton characteristics and accessibility for a broader spectrum of individuals with walking limitations.

## Data Availability

All data produced in the present study are available upon reasonable request to the authors

## Acknowledgments

Authors thank the clinical teams of the myelopathy program of CIUSSS-CN center and from the spinal cord injury programs of CMRRFK de Kerpape for their contribution to the completion of this study. Conducting interviews in France by N.F-B was made possible with travel grants received from the Canadian Institutes of Health Research CIHR, Mitacs Globalink program and the Bureau International of Université Laval.

