## Supplemental Material for "Acceptability of Overground Wearable Powered Exoskeletons for People with Spinal Cord Injury: a Multicenter Qualitative Study"

**Table S1. Inclusion and exclusion criteria for the use of the Indego® exoskeleton.**

| | |
|---|---|
| **Inclusion criteria** | |
| 1. | To be aged between 18 and 70 years old |
| 2. | To have an incomplete SCI |
| 3. | To be in the subacute stage of recovery (<1 year post-injury) |
| 4. | To be able to stand and/or walk therapeutically |
| 5. | To have a minimum/sufficient upper limb strength to use a rolling walker |
| 6. | To have a height between 5’1” and 6’1” |
| 7. | To have a weight <200 lbs (90 kg) |
| 8. | To have a length of femurs between 37 and 49 cm |
| 9. | To have a seated hip width < 42 cm |
| 10. | To have a medical approval to participate in the project following consultation with the multidisciplinary team |
| 11. | To have a standing tolerance >15 minutes |
| **Exclusion criteria:** Any medical condition or co-morbidity that may impair collaboration and participation or make wearing the exoskeleton unsafe, for example (non-exhaustive list): | |
| 1. | Having a walking speed > 0.6m/s |
| 2. | Disabling pain |
| 3. | Cognitive impairments affecting the ability to collaborate |
| 4. | Having skin lesions or wounds in the exoskeleton contact areas |
| 5. | Vertebral instability or spinal orthoses |
| 6. | Hip or knee contracture > 10° or ankle contracture > 5° |
| 7. | Severe or uncontrolled spasticity (score of 4 on the modified Ashworth scale) |
| 8. | Osteoporosis (criteria: T score < -3.0 for the proximal femur and/or BMD < 0.70g/cm2 for the distal femur and proximal tibia) or history of osteoporotic fractures |
| 9. | Non-healing fractures |
| 10. | Uncontrolled autonomic dysreflexia |
| 11. | Severe peripheral vascular disease |
| 12. | Severe heart failure |
| 13. | Active severe infection (abscess, osteomyelitis, Tuberculosis, etc.) |
| 14. | Lower limb prosthesis (amputation) and total knee replacement |
| 15. | Pregnancy |
| 16. | Colostomy bag |
| 17. | Active heterotopic ossification |
| 18. | Active deep vein thrombosis (untreated) |

**Table S2. Inclusion and exclusion criteria for the use of the Atalante® exoskeleton.**

| | |
|---|---|
| **Inclusion criteria** | |
| 1. | To be at least 18 years old |
| 2. | To have a paraplegia |
| 3. | To tolerate daily verticalization |
| 4. | To have a height between 160 cm and 190 cm |
| 5. | To have a weight <90 kg |
| 6. | To have a hip width less than or equal to 460 mm in a sitting position |
| 7. | To have thigh length between 380 and 460 mm |
| 8. | Leg length is also a criterion for inclusion, but this criterion varies according to user's range of motion in foot dorsiflexion. |
| **Exclusion criteria** | |
| 1. | History of osteoporotic fracture and/or disease or treatment leading to secondary osteoporosis with a threshold bone mineral density (BMD) value of 0.68 g/cm2. |
| 2. | Range of motion limitations below:<br>· For the hip: 115° flexion, 15° extension, 17° abduction, 10° adduction, 20° medial rotation, 10° lateral rotation.<br>· For the knee: 5° extension (flexum) and 110° flexion.<br>· For the ankle: 0° dorsiflexion (knee extended), 9° plantar flexion, 18° pronation and supination. |
| 3. | Severe spasticity in the adductor, hamstring, quadriceps, and triceps surae muscles (greater than 3 on the modified Ashworth scale). |
| 4. | Pregnant or breastfeeding women. |
| 5. | Decompensated psychiatric or cognitive disorders that could interfere with the proper use of the device. |
| 6. | History of osteoporotic fracture and/or disease or treatment leading to secondary osteoporosis. |
| 7. | Use of an active implantable device. |
| 8. | Evolving comorbid condition: pressure ulcers, infection, venous thrombosis. |
| 9. | Non-healing or unstable traumatic bone lesions in the limbs and pelvis. |
| 10. | Unstable spinal lesion. |
| 11. | Serious condition that could interfere with tolerance to exertion or the ability to perform static verticalization. |
| 12. | Evolving osteoma under medical advice. |
| 13. | Known syringomyelia under medical advice. |

**Interview questions.**

1. Before participating in the motorized exoskeleton walking training program, were you familiar with this approach? If yes, what did you know about it?

2. What would you have liked to know more about before starting to use an exoskeleton? For example, perceived benefits? Risks? What is the purpose of the exoskeleton?

3. Before starting the program, what problems did you anticipate regarding your participation in the exoskeleton walking training program?

4. Before starting the program, what were your hopes (hopes are a situation that will be ideal and preferred by an individual and will be influenced by their personal characteristics) for the motorized exoskeleton training program?

5. Before starting the program, what were your expectations (expectations concern a situation that is most likely to occur based on the individual's knowledge) for the motorized exoskeleton training program?

6. (Before starting) On a scale of 0 to 10, where 10 represents extremely and 0 not at all, how motivated were you to participate in the program/use the exoskeleton? For what reason(s) do you attribute this rating?

7. In your opinion, does starting the training program require particular skills? If so, what skills?

8. To continue and progress in the motorized exoskeleton training program, does it require particular [physical and cognitive] skills? If so, what skills?
    8a. Do you believe you have succeeded in acquiring these skills? Was it easy or difficult (complex)?

9. Can you think of ways that could have improved your competence or skills to perform the motorized exoskeleton training program/use the exoskeleton?

10. How easy or difficult is it to remember the steps involved in training with the exoskeleton?

11. To what extent does training with the exoskeleton required your attentional abilities (concentration)?

12. In your opinion, who should the motorized exoskeleton walking training program/the use of the exoskeleton be aimed at?

13. Why do you consider the training program appropriate for these individuals?

14. Do you believe that completing a walking program using a motorized exoskeleton could improve care for patients with spinal cord injury in the future?

15. Do you find it rather easy or rather difficult to participate in the motorized exoskeleton walking training program/use the exoskeleton? Why?

16. Currently (towards the end of training), are there any problems/difficulties that you perceive? Are there any steps in the program for which you are more or less confident in completing?

17. What, in your opinion, are the positive and negative consequences of your participation in the motorized exoskeleton walking training program?

18. Do you believe that the benefits of training with the motorized exoskeleton outweigh the efforts required?

19. If you had the opportunity, to what extent would you like to continue the motorized exoskeleton walking training program/the use of the exoskeleton?

20. (During training) On a scale of 0 to 10, where 10 represents extremely and 0 not at all, how motivated were you to continue the use of the exoskeleton? For what reason(s) do you attribute this rating?

21. Did you have specific goals when completing the motorized exoskeleton walking training program/the use of the exoskeleton?

22. During your training with the exoskeleton, were there elements in your environment that facilitated or limited your participation in the program/the use of the exoskeleton?
  - 22a. Schedule management?
  - 22b. The presence of the research team/therapists?
  - 22c. The location where the sessions took place?
  - 22d. The equipment?

23. How did you find the support and guidance from the research and/or clinical team?

24. What does your environment (family, friends, healthcare professional) think of the motorized exoskeleton walking training program/the use of the exoskeleton?

25. How do you feel about your participation in the motorized exoskeleton walking training program/the use of the exoskeleton (e.g., feelings)?

26. What is your overall satisfaction with the training sessions/the use of the exoskeleton? Very dissatisfied? Dissatisfied? Neither dissatisfied nor satisfied? Satisfied? Very satisfied?

27. Was the training with the exoskeleton relevant to you? Was the training adapted to your condition?

28. What did you like most about the training sessions?

29. What did you like least about the training sessions?

30. Do you have any other information you would like to share with us?
